# Genetic predisposition to loneliness increases schizophrenia and depression risk through inflammatory pathways: a Mendelian randomization study

**DOI:** 10.64898/2026.04.08.26350416

**Authors:** Coral I. Romualdo-Perez, Golam M. Khandaker, Eleanor Sanderson, Jennifer YF Lau, Livia A Carvalho

**Affiliations:** Translational Pharmacology Lab, William Harvey Research Institute, Charterhouse Square, Barts and the London School of Medicine and Dentistry, Queen Mary University of London, London, UK; Youth Resilience Unit, Wolfson Institute of Population Health, Queen Mary University of London, London, UK; MRC Integrative Epidemiology Unit, University of Bristol, Bristol, UK; Population Health Sciences, Bristol Medical School, University of Bristol, Bristol, UK; Centre for Academic Mental Health, Population Health Sciences, University of Bristol, Bristol, UK; NIHR Bristol Biomedical Research Centre, University Hospitals Bristol and Weston NHS Foundation Trust, Bristol, UK

## Abstract

**Background:** Loneliness is a psychosocial stressor associated with elevated risk of severe mental illness (SMI), including major depressive disorder (MDD), schizophrenia (SCZ), and bipolar disorder (BD). Loneliness is theorized to become biologically embedded via inflammation-related mechanisms, yet its causal relationship with SMI and the role of inflammatory signaling remain unclear.

**Aims:** To investigate whether loneliness causally influences SMI risk and whether inflammatory cytokines mediate this relationship.

**Method:** We applied univariable Mendelian randomization (MR) to estimate the causal effect of loneliness on SMI and multivariable MR (MVMR) to assess mediation by inflammatory signaling. We integrated genome-wide association study (GWAS) summary statistics for loneliness and SMI with genetic instruments for inflammatory cytokines. MVMR models estimated the direct effect of loneliness after accounting for inflammatory signaling using eQTL and pQTLs for interleukin-1 receptor antagonist (IL-1RA), interleukin-6 (IL-6), IL-6 receptor (IL-6R), tumor necrosis factor alpha (TNF-α), and TNF receptors (TNF-R1/2). Bidirectional MR examined potential reverse causal pathways between inflammation, SMI, and loneliness.

**Results:** MR provided evidence consistent with a causal effect of loneliness on SCZ and MDD. Results were also consistent with inflammatory cytokine pathways for IL-1RA, IL-6R, and TNF-R1, partially mediating the loneliness-SCZ and loneliness-MDD causal effect. No significant effects were identified for BD in UVMR or MVMR models. Bidirectional MR suggested evidence of reverse causation between SCZ and loneliness.

**Conclusions:** The findings support a causal risk-increasing effect of loneliness on SCZ and MDD, partially mediated by systemic inflammatory signaling, implicating pathways as a plausible mechanistic link between psychosocial stress and mental illness risk and highlighting potential opportunities for prevention and targeted intervention through inflammation and social pathways.

## 1. Introduction

### 1.1. Loneliness and severe mental illness

Loneliness is a chronic psychosocial stressor and an urgent global health issue. Chronic loneliness is associated with cognitive decline [1], decreased quality of life adjusted years [2], and an elevated mortality risk comparable to smoking or obesity [3, 4]. The increasing prevalence of loneliness across demographic groups, especially among socioeconomically disadvantaged populations, is heightening public health concerns [5, 6]. Additionally, current evidence demonstrates that loneliness is a heritable, transdiagnostic marker for poor mental health outcomes [7, 8], with established associations to mental disorders such as schizophrenia (SCZ) [9] and major depressive disorder (MDD) [10, 11].

### 1.2. Inflammation and biological mechanisms

Mounting evidence implicates inflammatory and immune dysfunction in both loneliness and severe mental illness (SMI). Loneliness has been associated with elevated interleukin-6 (IL-6) levels across adulthood [12], while meta-analyses identify increased IL-6 and IL-1 receptor antagonist (IL-1RA) as predictive markers for MDD [13] and SCZ [14], with broader inflammatory activation observed across mood disorders [15, 16]. Both chronic and acute stress further amplify systemic inflammation [17, 18], supporting a biological pathway linking loneliness to psychiatric risk via immune dysregulation; however, whether this reflects causality, reverse causation, or confounding remains unclear. Gene expression - an intermediate layer between genes and traits - varies across tissues and is influenced by expression quantitative trait loci (eQTLs), enabling tissue-specific evaluation of genetic effects and linking risk variants for the comorbidity of social isolation and SMI to underlying pathology.

### 1.3. Study rationale and aims

To better understand how social stressors are biologically embedded in SMI risk, we tested the hypothesis that inflammation mediates the causal effect of loneliness on SMI outcomes. The Social Signal Transduction Theory of SMI [19] conceptualizes social stressors, such as loneliness, as activators of inflammatory pathways, yet no prior study to date has jointly modeled loneliness and cytokines within a causal framework; although cytokines suggest causal and therapeutic potential [20, 21], studies examining whether inflammation mediates the causal effect between psychosocial stress and psychiatric risk remains limited, despite its potential to identify mechanistic pathways linking loneliness, SMI, and treatment targets.

To address this gap, we used Mendelian randomization (MR), which leverages genetic variants as instrumental variables to estimate causal effects while reducing confounding and reverse causation [22], alongside multivariable MR (MVMR) to estimate direct effects of correlated exposures [23]. We hypothesized that inflammation mediates the association between loneliness and SMI; first testing the causal effect of loneliness on SCZ, MDD, and BD using MR, followed by MVMR to assess inflammatory mediation [24], and bidirectional analyses using eQTL and pQTL data. These insights may inform diversity and inclusion policies by clarifying why SMI is more prevalent in populations facing chronic loneliness, why it frequently co-occurs with mental health conditions, and why it often recurs. This work may also suggest new opportunities for preventing and treating SMI by targeting inflammation.

## 2. Methods

### 2.1. Study Design

This study, conducted as part of the MSc in Genomic Medicine at QMUL by CIRP [25], employed a two-sample, summary-data Mendelian randomization (MR) design to investigate whether loneliness affects the risk of severe mental illness (SMI) through inflammatory signaling. MR assumes (i) the genetic instruments are associated with the exposure (relevance), (ii) the instruments are not associated with exposure–outcome confounders (independence), and (iii) the instruments affect the outcome only through the exposure (exclusion restriction) [26]. We applied both univariable MR (UVMR) and multivariable MR (MVMR) [24, 27] (Fig. 1), where UVMR estimated total effects and MVMR jointly modeled loneliness and cytokine traits to estimate direct effects and partial mediation of the loneliness–SMI association. Analyses incorporated genetic instruments for loneliness and cis-instruments for cytokines, alongside pairwise bidirectional UVMR and MVMR analyses across loneliness, cytokines, and SMI. We used de-identified publicly available GWAS and QTL summary statistics from predominantly European ancestry cohorts (Fig. 2). No new data was collected, and no individual-level data was accessed. Therefore, no additional ethical approval was required for this study.

**Figure 1.**
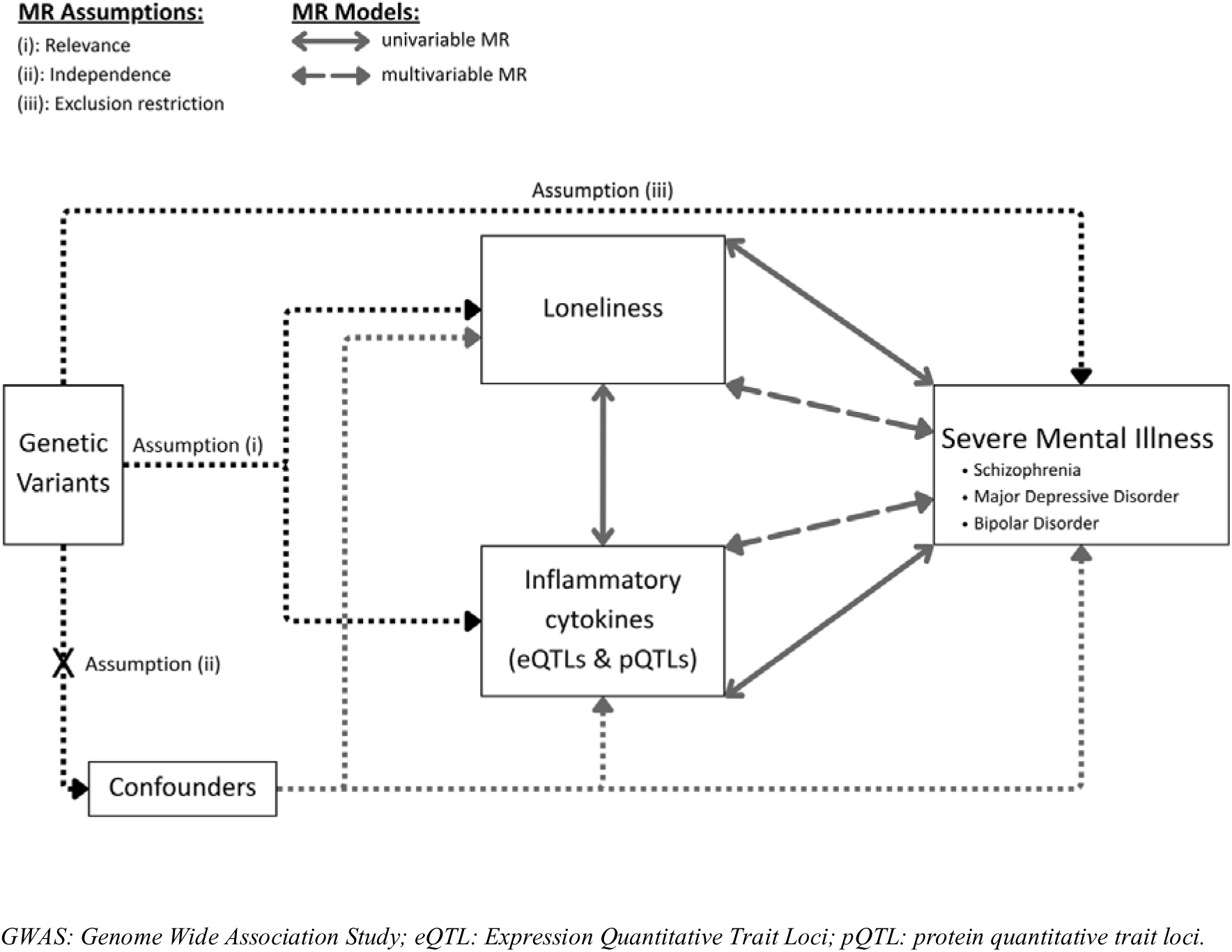
Conceptual framework for Mendelian randomization analyses of loneliness, inflammatory cytokines, and severe mental illness. Diagram illustrating the hypothesized causal effects tested using univariable and multivariable Mendelian randomization (MR). Genetic variants serve as instrumental variables (IVs) associated with loneliness (GWAS) and inflammatory cytokine levels (eQTLs and pQTLs). Univariable MR evaluates the effect of loneliness and inflammatory cytokines on severe mental illness (schizophrenia, major depressive disorder, and bipolar disorder) as well as the bidirectional relationship between loneliness and inflammation. Multivariable MR estimates the direct effects accounting for inflammation as a potential mediator.

**Figure 2.**
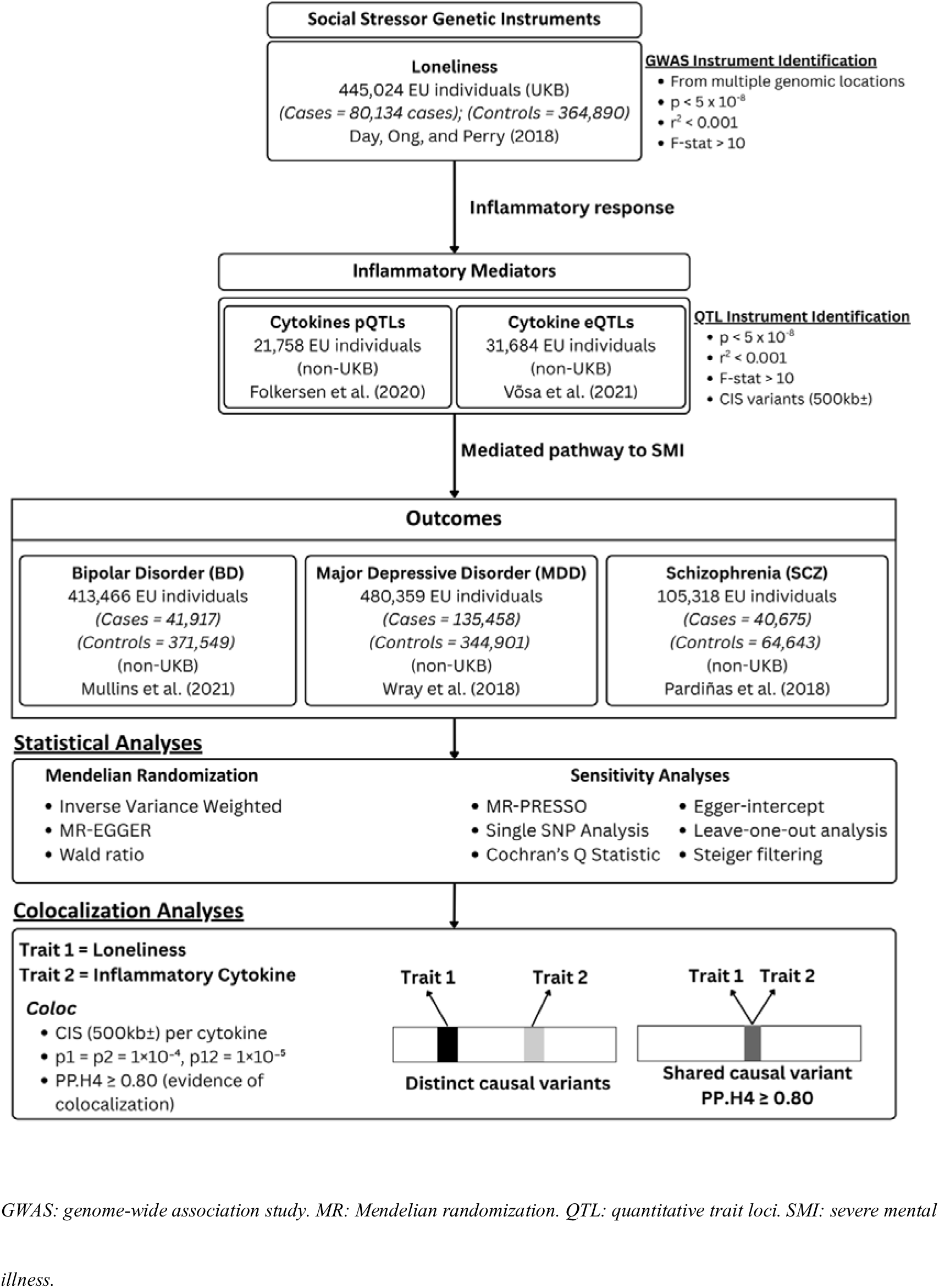
Flow analysis. Data sources, instrument selection strategy, and statistical analysis pipeline for Mendelian randomization.

### 2.2. Data Sources

Genetic instruments for loneliness were obtained from a GWAS by Day, Ong, and Perry (2018) in 452,302 UK Biobank participants of European ancestry, see Fig. 2. Loneliness phenotype was defined using responses to the question, “Do you often feel lonely?” Participants answering “Yes” were cases (80,134) and those answering “No” were controls (364,890).

Blood eQTL summary statistics for seven inflammatory cytokine genes – interleukin-1 beta (IL-1β), interleukin-1 receptor antagonist (IL-1RA), interleukin-6 (IL-6), interleukin-6 receptor (IL-6R), tumor necrosis factor alpha (TNF-α), TNF receptor 1 (TNF-R1), and TNF receptor 2 (TNF-R2) – were extracted from eQTL Gen Phase 1 (n = 31,684) [29]. Blood pQTL summary statistics, n = 21,758, for IL-1RA, IL-6, IL-6Rα, TNF-R1, and TNF-R2 were obtained from Folkersen et al. (2020). We analyzed plasma eQTL and pQTL datasets separately to account for platform- and measurement-specific differences.

Outcome GWAS were restricted to European ancestry samples to minimize population stratification (Fig. 2). To reduce bias from sample overlap with the UK Biobank, psychiatric outcomes were sourced from non-UK Biobank Psychiatric Genomics Consortium datasets: schizophrenia (SCZ; CLOZUK; 40,675 cases and 64,643 controls) [31], major depressive disorder (MDD; 135,458 cases and 344,901 controls) [32], bipolar disorder (BD; 41,917 cases and 371,549 controls) [33]. For replication, we used two independent plasma proteomic resources from For replication, we used two independent plasma proteomic resources from Ferkingstad et al., 2021 (n = 35,559) and UK Biobank Pharma Proteomics Project (n ≈ 54,000) [35] and a cerebrospinal fluid (CSF) proteomic dataset from Western et al., 2024 (n = 3,506). Where required, we used UCSC liftOver to convert GRCh38 coordinates to GRCh37 [37, 38] and assess consistency of effect direction, effect size, and instrument strength across datasets.

### 2.3. Instrument Selection and Harmonization

For loneliness, cytokine QTLs, and psychiatric outcomes, we selected genome-wide significant SNPs (p < 5×10□□) and performed LD clumping (r² < 0.001) using the European 1000 Genomes reference panel [39] via OpenGWAS [40]. Cytokine instruments were restricted to cis variants (±500 kb), with additional conditionally independent SNPs identified using GCTA-COJO where available [41]. Data were harmonized to the same effect allele using TwoSampleMR (v0.6.29) [42], and proxy SNPs (r² ≥ 0.8) were identified using LdlinkR (v1.4.0) when needed [43]. Instrument strength was assessed using mean F-statistics (UVMR) and conditional F-statistics (MVMR), with F > 10 indicating strong instruments.

### 2.4. Univariable Mendelian Randomization Analyses

We used inverse-variance weighted (IVW) MR as the primary estimator and the Wald ratio for single-SNP instruments. In a bidirectional framework, we tested total effects between loneliness, cytokine traits, and SMI outcomes (SCZ, MDD, BD), including all pairwise directions. Robustness assessments (≥3 SNPs) included MR-Egger, weighted median, and weighted mode. Primary analyses used full instrument sets and were repeated after excluding variants contributing to heterogeneity or pleiotropy. Heterogeneity and pleiotropy were assessed using Cochran’s Q and the MR-Egger intercept, respectively, with MR-PRESSO used to detect and correct pleiotropic outliers. MR Steiger filtering was applied to limit reverse causation, and leave-one-out and single-SNP analyses identified influential variants. For loneliness instruments (N = 13), sensitivity analyses removed 5 variants for SCZ (including 2 overlapping between Steiger and MR-PRESSO), and 1 and 2 variants for MDD and BD, respectively, identified through MR-PRESSO (Tables S3-S4).

### 2.5. Multivariable Mendelian Randomization Analyses

We applied Multivariable Mendelian randomization (MVMR) [23] to estimate the direct effects of loneliness and cytokine traits on SMI outcomes, accounting for shared genetic architecture [26, 43], using instruments for loneliness and cis cytokine variants in a combined model. Directionality was further assessed using bidirectional models incorporating SMI instruments with cis cytokine instruments and loneliness as the outcome. Analyses were conducted in R (multivariable MR package v0.4) using multivariable IVW regression as the primary estimator, with conditional F-statistics for instrument strength [43], multivariable MR-Egger for pleiotropy [44–45], and Q-statistics for residual heterogeneity [23].

All analyses used a two-sample framework assuming independent samples. For binary traits (loneliness, MDD, SCZ, BD), estimates (beta, OR, 95% CI) were scaled to reflect a one standard deviation increase in genetic liability using 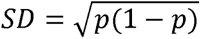, where *p* was the case proportion in the source GWAS, while QTL exposures were retained in their original SD scale. Results are reported as beta coefficients, standard errors, and p-values, with only statistically significant findings carried forward to colocalization analyses (Fig. 1).

### 2.6. Colocalization Analyses

For exposure–outcome pairs with significant MR and MVMR estimates, we used Bayesian colocalization to assess whether significant associations between loneliness and SMI reflect shared causal variants [46, 47], extracting ±500 kb (and ±250 kb sensitivity) regions around loneliness lead SNPs, SCZ loci, and cytokine genes with significant effects. Analyses were conducted using coloc (v5.1.0) with priors p1 = p2 = 1×10□□ and p12 = 1×10□□, defining evidence of colocalization as PP.H4 ≥ 0.80 and sweeping sensitivity analyses across p12 (10□□–10□²). Results were interpreted cautiously due to reliance on detectable regional signals, as limited power can hinder distinction between shared (H4) and distinct (H3) variants [48, 49].

### 2.7 Interpretation Framework: Tier Classification

Because psychosocial traits can yield weaker instruments (i.e., lower correlation with the exposure), we summarized exposure using a tiered framework (Table S2) based on statistical evidence, instrument strength, and diagnostic checks for heterogeneity and pleiotropy. Tier A required stronger statistical evidence (P < 0.01), F-statistics > 10, and consistent sensitivity results. Tier B required P < 0.05 with strong instruments (F > 10) and directional support; Tier B* indicates Tier B results with a single, unresolved pleiotropy flag. Tier C captured weaker instruments (F < 10) or incomplete support, and Tier D reflected evidence of heterogeneity or pleiotropy across multiple diagnostics, low instrument strength, and a lack of directional support. This framework was used to aid interpretation rather than as a formal grading system.

## 3. Results

### 3.1. Univariable MR: Is loneliness likely causal to severe mental illness?

To confirm directionality, we first assessed whether loneliness had a causal effect on SMI through UVMR. Loneliness was identified to have a risk-increasing potential causal effect on SCZ (IVW p < 0.05), see Fig. 3, validated by other MR estimates, and MDD (IVW p < 0.05), validated by all other MR methods except MR-EGGER (Fig. 4). Potential horizontal pleiotropy was identified in both analyses through MR-PRESSO (p < 0.05) with no other outlier indices (Table S3). No significant effects were observed for loneliness on BD.

**Figure 3.**
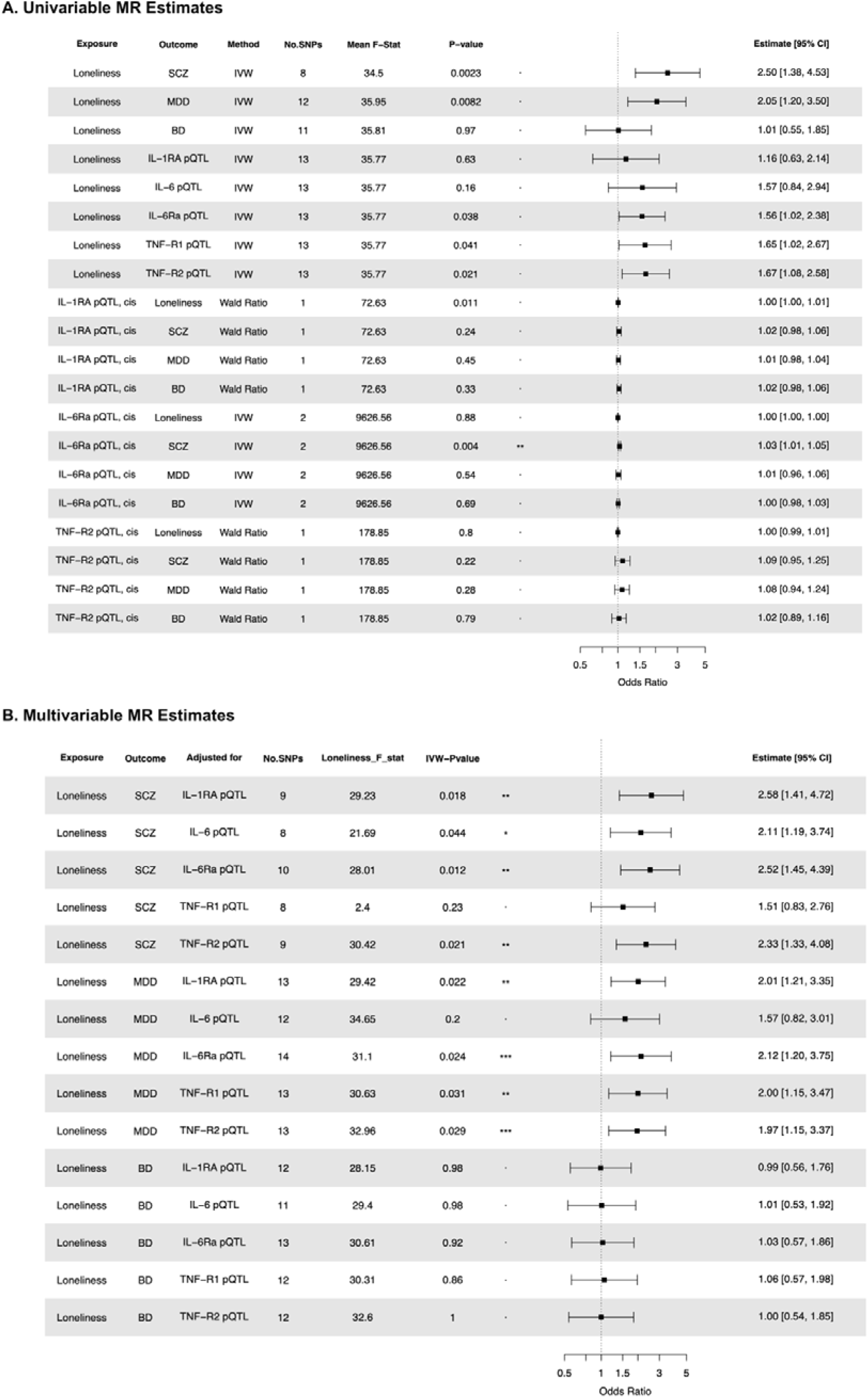
Multivariable MR and univariable MR Estimates of Loneliness instruments and Inflammatory Cytokine on Severe Mental Illness. Estimates assess the direct and total effects of loneliness and cytokine pQTLs on schizophrenia (SCZ), major depressive disorder (MDD), and bipolar disorder (BD). (*) = significant results validated in one dataset. (**) = significant results validated in two datasets. (***) = significant results validated in three datasets. Plots show odds ratio (OR) with 95% CIs on 1-SD of the exposure.

**Figure 4:**
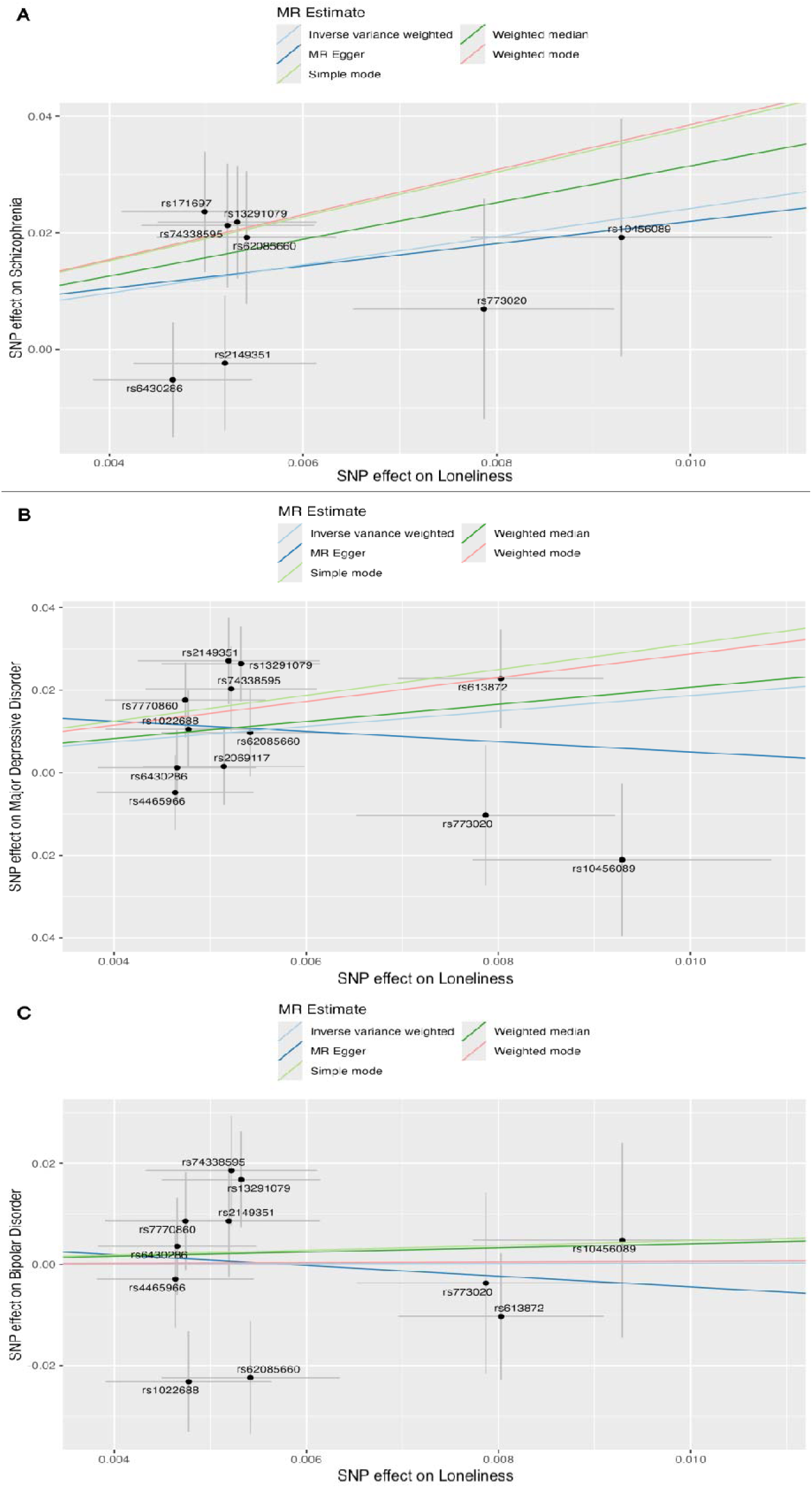
Scatterplot of MR Estimates for Loneliness to Severe Mental Illness outcomes using instruments retained after sensitivity assessments. Panel A: schizophrenia. Panel B: major depressive disorder. Panel C: bipolar disorder.

Initial analyses using the complete loneliness instrument list, which indicated heterogeneity and pleiotropy, showed a significant causal effect but with conflicting beta directions observed between IVW and MR-EGGER (Tables S3-S4). Further, sensitivity assessments revealed significant heterogeneity (Cochran’s Q, p < 0.01) and horizontal pleiotropy (MR-Egger intercept, p < 0.01; MR-PRESSO, outlier test p < 0.01) with several variants identified as outliers (Table S3). We found that although the MR-PRESSO distortion test was non-significant for both MDD and SCZ (Table S5), the identification of outlier SNPs (outlier p < 0.05) indicated that key instruments may violate MR assumptions of (ii) independence and (iii) exclusion due to pleiotropic instruments.

### 3.2. Univariable MR: Does loneliness causally influence immune dysregulation?

Next, we assessed whether loneliness had a causal effect on circulating inflammatory cytokine levels. UVMR analyses indicated that loneliness had a risk-increasing potential causal effect on IL-6Rα_pQTL_, TNF-R1_pQTL_, and TNF-R2_pQTL_, see Table S5. These effects were not observed in additional analyses using cytokine pQTLs from plasma and CSF. Sensitivity assessments showed no evidence of heterogeneity or pleiotropy (Table S4), aside from loneliness to TNF-R2_pQTL_ via MR-PRESSO (p = 0.039; no outliers). No significant results were observed for loneliness on IL-1RA_pQTL_ or IL-6_pQTL_ across all estimates. Analyses assessing the direct effects of loneliness on cytokine cis-eQTLs could not be performed due to a lack of instrumental overlap.

### 3.3. Univariable MR: Is immune dysregulation causally associated with loneliness or severe mental illness?

Following, we assessed whether cis-QTL instruments for cytokines had a significant effect on loneliness or SMI. This showed that IL-1RA_pQTL_ had a potentially causal risk-increasing effect on loneliness, but this was not replicated with cis-pQTL instruments obtained from validation datasets (Table S3). Sensitivity assessments could not be performed for this model due to only one instrument being available (Table S4). No other significant effects were observed for IL-6Rα_pQTL_ and TNF-R2_pQTL_ on loneliness.

Assessment of total effects of inflammation on SMI showed a positive causal effect of IL-6Rα_pQTL_ on SCZ, with no evidence of heterogeneity or pleiotropy (Table S4) and replication in validation datasets (Table S3). No other significant effects (IVW p < 0.05) were identified for IL-1RA_pQTL_, IL-6Rα_pQTL_, and TNF-R2_pQTL_ on MDD or BD; assessment of the presence of heterogeneity and pleiotropy could not be performed for cytokines with a single SNP available (Table S4). No instruments were available for IL-6_pQTL_ or TNF-R1_pQTL_. No significant effects were observed for any of the cis-eQTLs (Wald ratio p > 0.05) on loneliness or SMI outcomes (Table S3).

### 3.4. Univariable MR: Does severe mental illness causally influence loneliness or immune dysregulation?

We further assessed whether the causal effects identified above were bidirectional. A significant positive causal effect from SCZ to loneliness (IVW p < 0.01) was identified. However, directional pleiotropy (MR-EGGER p < 0.01) and horizontal pleiotropy (MR-PRESSO p < 0.01; no outliers) remained significant with no further outliers. Initial results did not meet MR assumptions due to significant heterogeneity (Cochran’s Q < 0.05), directional pleiotropy (MR-EGGER p < 0.005), and horizontal pleiotropy (MR-PRESSO p < 0.05), with several SNPs identified as outliers (Table S3-S4). Assessments of MDD and BD on loneliness were both null across all estimates.

Further analyses assessed whether SMI had causal effects on circulating inflammatory cytokine levels. Within these results, no significant effects (IVW p > 0.05) were observed for SCZ or MDD on IL-1RA_pQTL_, IL-6_pQTL_, IL-6Rα_pQTL_, TNF-R1_pQTL_, or TNF-R2_pQTL_ (Table S3). A causal, negative effect of BD on TNF-R1 was identified with validation in the CSF dataset; potential horizontal pleiotropy was identified through MR-PRESSO (p < 0.05) with no significant outliers (Table S4).

### 3.5. Multivariable MR: Does inflammation mediate the association between loneliness and severe mental illness?

We assessed whether inflammation partially mediated the causal effect of loneliness on SMI. A risk-increasing potential causal effect of loneliness on SCZ was observed after adjusting for IL-1RA_pQTL_ (Tier B*), IL-6Rα_pQTL_ (Tier B*), and TNF-R2_pQTL_ (Tier D) (Fig. 3). We could not reject the null hypothesis of no heterogeneity (Cochran’s Q, p > 0.05). Similar causal effects of loneliness on MDD were observed after adjusting for IL-1RA_pQTL_ (Tier B*), IL-6Rα_pQTL_ (Tier D), and TNF-R2_pQTL_ (Tier D). The aforementioned inflammatory pathways, both for SCZ and MDD, were further validated in secondary cytokine pQTLs from plasma, showing consistent partial mediation of the direct effect of loneliness on SCZ and MDD (Table S6). Initial analyses using the complete loneliness set – suggested the presence of heterogeneity and pleiotropy – were only significant for MDD (Tables S6-S7). As no cis-instruments were available for IL-6_pQTL_ and TNF-R1_pQTL_, these cytokines could not be incorporated into the analyses. No significant results were observed for BD (Fig. 3).

### 3.6. Multivariable MR Results: Does inflammation mediate the association between severe mental illness and loneliness?

Next, we assessed whether the causal effect above was bidirectional; inflammation was a potential mediator between SMI and loneliness. A risk-increasing potential causal effect (Table S6) of SCZ on loneliness was observed after adjusting for IL-1RA_pQTL_ (Tier D), IL-6Rα_pQTL_ (Tier D), and TNF-R2_pQTL_ (Tier D). However, there was significant heterogeneity in all models (Table S7). For MDD, no causal effect on loneliness was observed after adjusting for IL-6RαpQTL; the other models were not estimable due to a lack of sufficient instruments (SNPs ≤ 2; Table S6). No significant effects were observed utilizing instruments for BD and cis-pQTLs on loneliness.

### 3.7. Colocalization: Is there a shared causal variant between loneliness, schizophrenia, and immune dysregulation?

Colocalization analyses at loneliness-associated loci assessed shared causal variants between loneliness, SCZ, MDD, and inflammatory mediators (IL-1RA, IL-6R, TNF-R2). For SCZ, loneliness-led analyses favored loneliness-only (H1) or distinct variants (H3), with two loci supporting H3 (PP.H3 = 0.85–0.88; Table S8) and no changes across sensitivity analyses (Table S9). SCZ-led analyses similarly favored SCZ-only models (PP.H2 > 0.80), with two loci supporting H3 (PP.H3 ≈ 0.99) and one supporting colocalization (PP.H4 ≈ 0.95; Table S8), unchanged in sensitivity analyses (Table S9) and ±250 kb windows supporting H1/H2.

For MDD, loneliness-led analyses primarily favored H1, but sensitivity analyses identified one locus (rs13291079) supporting colocalization (PP.H4 ≈ 0.98; Table S8). MDD-led analyses similarly identified one colocalized locus (rs76025409; PP.H4 ≈ 0.94), with remaining loci favoring MDD-only models (H2; Table S9) and no changes across sensitivity or ±250 kb analyses (Tables S8-S9).

Next, we assessed inflammation-led loci, which predominantly favored pQTL-only models (H1) across IL-1RA, IL-6, IL-6R, TNF-R1, and TNF-R2 [27, 31, 32] (Tables S10- S11), with limited evidence of colocalization except IL-1RA (PP.H4 = 0.85; 0.81 in sensitivity analyses). Inflammation–SCZ analyses primarily supported H1 with H3 support within IL-6R (Table S10), while inflammation–MDD analyses consistently favored H1 with no changes in sensitivity or a ±250 kb window. Overall, these results support distinct causal variants at IL-6R for SCZ and no colocalization with loneliness under current power, with limited support for MDD at single loci in each direction.

## 4. Discussion

### 4.1. Summary of Main Findings

Across our Mendelian randomization analyses, we observed genetic evidence supporting an effect of loneliness on severe mental illness (SMI), with pro-inflammatory signaling as a plausible mediator. Findings were consistent for schizophrenia (SCZ) and major depressive disorder (MDD), where loneliness showed total and direct effects, and circulating IL-1RA, IL-6Rα, and TNF-R2 were consistent with partial mediation and risk contribution, whereas bipolar disorder (BD) showed only a total effect on TNF-R1 without other robust associations. Colocalization analyses identified distinct causal variants at two loci between loneliness and SCZ and one potential shared variant between loneliness and MDD. There was no cis-colocalization between loneliness and IL-1RA, IL-6/IL-6R, or TNF-R1/2 across panels, but there was evidence of distinct variants between IL-6R and SCZ. Overall, these results support disorder-specific immune mechanisms, with IL-1RA, IL-6Rα, and TNF-R2 as plausible partial mediators for SCZ and MDD, limited evidence for BD, and inflammation emerging as a candidate pathway linking loneliness to increased risk of SCZ and MDD.

### 4.2. Schizophrenia: Immune Pathways Linking Loneliness to Psychosis

Our findings provide genetic evidence consistent with loneliness increasing schizophrenia (SCZ) risk through multiple inflammatory pathways. Loneliness instruments adjusted for IL-1RA expression retained a causal effect with limited evidence of pleiotropy or heterogeneity, supporting IL-1RA as a partial mediator, aligning with evidence that elevated IL-1RA reflects chronic immune activation or compensatory anti-inflammatory signaling in SCZ [50–52]. IL-6Rα protein levels also indicated partial mediation, aligning with prior MR studies implicating IL-6 signaling in SCZ [53], with soluble IL-6R trans-signaling driving pro-inflammatory responses linked to neuroinflammation, stress reactivity, and impaired neurogenesis [54–55]. TNF-R2 levels also increased the loneliness–SCZ effect estimate when modeled jointly, consistent with partial mediation, with TNF-R2 implicated in neuroprotection [56] and SCZ severity [57–58]. Replication across independent datasets for IL-1RA, IL-6Rα, and TNF-R2 provides triangulated evidence supporting a causal role, collectively implicating IL-1RA, IL-6R, and TNF pathways in the biological embedding of loneliness within SCZ risk.

### 4.3. MDD: Immune Pathways Linking Loneliness to Depression

Our analyses provide genetic evidence consistent with loneliness influencing major depressive disorder (MDD) risk, in line with prior work linking loneliness to depression via social stress pathways. Loneliness adjusted for IL-1RA levels retained a causal effect with limited heterogeneity and pleiotropy, consistent with observational and genetic evidence implicating IL-1RA in depression [59–61]. IL-6Rα protein levels similarly supported partial mediation, with replication across plasma and cerebrospinal fluid and prior evidence linking soluble IL-6R [62] and IL-6 signaling to increased depression risk [63]. TNF-R2 showed comparable effects, supported by animal models where TNF-R1/2 deletion induces antidepressant-like responses [64] and elevated levels observed in severe mental illness, including depression [58]. Colocalization with sensitivity analyses identified a shared causal variant (rs13291079) between loneliness and MDD; this SNP lies within an intron of PHF2 [65–67] and has been shown to act as a co-activator with NF-κB in macrophages, promoting pro-inflammatory responses [68]. Overall, these findings support inflammatory pathways as mediators linking loneliness to depression risk.

### 4.4 BD: Limited Genetic Evidence for Inflammatory Mediation

Further, evidence for BD was limited. A single MR analysis suggested a potential negative effect of BD on TNF-R1pQTL, contrasting with previous reports linking BD with increased TNF-R1 levels [57, 69]. The overall pattern of null results may reflect reduced power and etiological heterogeneity within BD, including subtype differences (e.g., higher SNP-based heritability in BD I compared with BD II) [70, 71] alongside complex polygenic architecture [72]. Future work using larger, stratified, and multi-ancestry GWAS, together with improved inflammatory QTL instruments, will be needed to clarify whether loneliness and inflammatory signaling play distinct roles in BD pathophysiology.

### 4.5. Limitations

Despite integrating eQTL and pQTL instruments to enhance interpretability, several limitations remain. Residual heterogeneity and pleiotropy cannot be fully excluded, although sensitivity analyses generally support MR assumptions, and limited statistical power — particularly for loneliness and exposures with low SNP counts or F-statistics < 10 — may introduce weak instrument bias and inflate estimates, warranting careful interpretation of results, as it may cause inflated causal effect estimates. Some cytokines (e.g., IL-6pQTL and TNF-R1pQTL) lacked sufficient instruments for MVMR, and protein data for TNF-α and IL-1β were unavailable, while discordance between IL-6Rα protein and full IL-6R expression may reflect bias due to exclusion of the beta subunit (unavailable in source data). Further, social traits such as loneliness remain vulnerable to confounding from population structure and dynastic effects [73], and restriction to European-ancestry cohorts limits generalizability.

### 4.6. Future Directions

Future work should examine loneliness–inflammation effects at the symptom or dimensional level, incorporate trans-ancestry analyses, and leverage larger GWAS of social traits alongside expanded, tissue-specific pQTL resources to improve instrument strength. If validated by complementary approaches, these findings may inform prevention and intervention strategies targeting both social isolation and inflammation within individuals.

### 4.7 Conclusions

In summary, our findings provide genetic evidence consistent with loneliness increasing risk for schizophrenia (SCZ) and major depressive disorder (MDD), with inflammatory signaling—particularly IL-1RA, IL-6R, and TNF pathways—acting as plausible partial mediators. Our use of multivariable MR enabled the separation of direct and indirect effects, highlighting immune mechanisms underlying disorder-specific vulnerability. These results support a model in which psychosocial stress becomes biologically embedded through immune mechanisms, contributing to disorder-specific vulnerability. While evidence for bipolar disorder (BD) was limited, the overall pattern highlights inflammation as a potential pathway linking loneliness to psychiatric risk. If validated in future studies, these findings may inform prevention and intervention strategies targeting both social and biological determinants of severe mental illness.

## Data Availability

All data produced are available online through Zenodo (10.5281/zenodo.18473530); this includes all analytical code and study-derived outputs,

https://www.doi.org/10.5281/zenodo.18473530

## Disclosures and Declarations of Interest

LAC has received funding and consultancy fees from MINDLIFE. LAC and GMK have no conflict of interest to declare with regard to the content of this manuscript. For full disclosure, LAC has received consultancy fees from MINDLIFE and King Faisal Specialist Hospital & Research Centre; GMK has received royalties from the Cambridge University Press for the Textbook of Immunopsychiatry, and consultancy fees from the Neuroimmune Foundation and the Danish Research Fund (DFF). All other authors report no financial relationships with commercial interests. Claude (Anthropic, claude.ai, model: Claude Opus 4, accessed January–March 2026) assisted in the language and flow of the manuscript.

## Funding

LAC is supported by the Wellcome Trust (226777/Z/22/Z), Barts Charity (G001414), and the UKRI Social Health Hub of the Mental Health Platform (MR/Z503514/1). JL is supported by the UKRI Social Health Hub of the Mental Health Platform (MR/Z503514/1). GMK is supported by the UK Medical Research Council (MRC; MC_UU_00032/6), which contributes to the MRC Integrative Epidemiology Unit at the University of Bristol, and by the Wellcome Trust (201486/Z/16/Z and 201486/B/16/Z), the MRC (MR/W014416/1, MR/S157675/1, MR/Z50354X/1, and MR/Z503745/1), and the National Institute for Health and Care Research Bristol Biomedical Research Centre (NIHR 203315). ES is supported by UKRI (UKRI0077) and the European Union (101073237; ESSGN). The views expressed in this publication are those of the authors and do not necessarily reflect those of the European Union, MSCA Horizon Europe,, ESSGN, NIHR, or the Department of Health and Social Care.

## Author contributions

LAC: Conceptualisation, Methodology, Writing – Original Draft Reviewing & Editing

CRP: Conceptualisation, Methodology, Formal Analysis, Original draft – Reviewing & Editing

GK: Methodology, Writing – Reviewing & Editing

ES: Methodology, Writing – Reviewing & Editing

JL: Reviewing & Editing

## Data and Analytical Code Availability

All data used in this study are publicly available. Summary statistics for perceived social isolation (PSI; loneliness/social isolation traits) were obtained from Day et al. (UK Biobank) via the NHGRI-EBI GWAS Catalog under accession number (https://www.ebi.ac.uk/gwas/studies/GCST006923). Expression summary data was obtained from Võsa et al. via eQTL Gen Phase 1. Plasma protein pQTL summary data were taken from Folkersen et al. via GWAS catalog (IL-1RA <u>GCST90012004</u>; IL-6 <u>GCST90012005</u>; IL-6Rα <u>GCST90012025</u>; TNF-R1 <u>GCST90012015</u>; TNF-R2 <u>GCST90012026</u>). Proteomic validation datasets for Ferkingstad et al. were accessed via the deCODE public summary-data portal (https://www.decode.com/summarydata), and the UK Biobank Pharma Proteomics Project (UKB-PPP) pQTL data were obtained from the Sun et al. repository (http://ukb-ppp.gwas.eu). Summary statistics used for severe mental illness (SMI) endpoints were accessed through the Psychiatric Genomics Consortium (PGC) and are publicly available from the PGC downloads portal under the relevant disorder workgroups. Analytical code, study-derived outputs, and result tables are provided in the Supplementary Data and through Zenodo (<u>10.5281/zenodo.18473530</u>).

## Notes

### Author Declarations

All data used in this study are publicly available. Summary statistics for perceived social isolation (PSI; loneliness/social isolation traits) were obtained from Day et al. (UK Biobank) via the NHGRI-EBI GWAS Catalog under accession number (https://www.ebi.ac.uk/gwas/studies/GCST006923). Expression summary data was obtained from Vosa et al. via eQTL Gen Phase 1. Plasma protein pQTL summary data were taken from Folkersen et al. via GWAS catalog (IL-1RA GCST90012004; IL-6 GCST90012005; IL-6Rα GCST90012025; TNF-R1 GCST90012015; TNF-R2 GCST90012026). Proteomic validation datasets for Ferkingstad et al. were accessed via the deCODE public summary-data portal (https://www.decode.com/summarydata), and the UK Biobank Pharma Proteomics Project (UKB-PPP) pQTL data were obtained from the Sun et al. repository (http://ukb-ppp.gwas.eu). Summary statistics used for severe mental illness (SMI) endpoints were accessed through the Psychiatric Genomics Consortium (PGC) and are publicly available from the PGC downloads portal under the relevant disorder workgroups.

### Summary of Updates

Author order; supplemental table order and files updated.

